# Early experiences with antibody testing in a Flemish nursing home during an acute COVID-19 outbreak: a retrospective cohort study

**DOI:** 10.1101/2020.05.18.20105874

**Authors:** Buntinx Frank, Claes Peter, Gulikers Marjo, Jan Y. Verbakel, De Lepeleire Jan, Michaël C.J. Van der Elst, Marc Van Ranst, Pieter Vermeersch

**Author notes:** Correspondence: Michaël Van der Elst, Dept, of Public Health and Primary Care, KU Leuven, Leuven, Belgium.

## Abstract

**objectives:** to assess the prevalence of COVID-19 (PCR-test) in residents and staff of a nursing home. To examine the presence of IgM and IgG antibodies in the sample and the relation between PCR and antibody test results.

**design:** cross-sectional and (retrospective) cohort study

**setting:** a nursing home for the elderly Bessemerberg in Lanaken (Belgium) with up to 130 beds. Lanaken is situated in the Belgian province with the highest COVID-19 prevalence.

**participants:** residents (N=108) and staff members (N=93) of the nursing home

**outcomes:** PCR, IgM and IgG

**results:** the prevalence of COVID-19, based on PCR test was 34% (N=40) for residents and 13% (N=11) for staff members, respectively. Of the residents, 13% showed positive IgM results and 15% positive IgG results. In 17% of the residents, at least one of the antibodies was positive. In total 13% of the staff members had positive IgM and 16% had a positive IgG. In 20% of the staff members at least one of these antibody tests was positive. In PCR positive residents, the percentage of IgM positive, IgG positive, and at least one of both was 28%, 34%, and 41%. In PCR positive staff, we found 30%, 60%, and 60%. Additional antibody tests were performed in nine residents between day 11 and 14 after the positive PCR test. Of those, 7 (78%) tested positive on at least one antibody. When retesting three weeks later, all remaining residents also tested positive.

**conclusions:** Recently it was reported that in Belgium antibodies are present in 3-4% of the general population. Although, the prevalence in our residents is higher, the number is largely insufficient for herd immunity. In staff members of the regional hospital the prevalence of antibodies was 6%. The higher prevalence in nursing home staff (21%) may be related to the complete absence of good quality protection in the first weeks of the outbreak.

**Article summary:**

Strengths and limitations of this study
- This is the first study in Belgium examining the prevalence of COVID-19 and the presence of antibodies in residents and staff members of a nursing home
- The internal procedural control was positive -with one exception- in all tests, which suggests good quality sampling and testing.
- Some degree of selection bias should be assumed in residents, since some residents were absent; mostly from hospitalisation or death which can be related to the presence of COVID-related disease.
- The study was set up in one nursing home and is consequently not representative for the whole of the Flemish community

## Introduction

On 31 December 2019, a cluster of pneumonia cases of unknown aetiology was reported in Wuhan, Hubei Province, China.[1] Since then, the SARS-CoV-2 virus causing the severe acute respiratory syndrome called coronavirus disease (COVID-19), succeeded to spread across the world and has been entitled as a pandemic.[2] The first confirmed case of a SARS-CoV-2 infection in Belgium, identified on February 4^th^ was a businessman returning from China who was successfully quarantined in the following weeks.[3] The second positive case was documented on February 29^th^, followed by a series of new positive cases and confirmed local transmission early March 2020. Afterwards the virus was rapidly spread throughout the entire country leading to instigated rules by the government.

In Belgium, the SARS-CoV-2 rapidly spread within medical and nursing facilities. Currently, the majority of the people whom died of COVID-19 occurred in nursing homes, and not in a hospital setting.[4] Thereby, a shortage of health workers in these nursing home facilities is at risk due to the infection of many staff members; which may put further pressure on the already strained health systems.

The nursing home for the elderly (woon- en zorgcentrum (WZC)) Bessemerberg in Lanaken (Belgium) is a private long-term care facility, founded in 2011 with a normal capacity of 130 beds. Mean age of the residents is 86 years. It is worthwhile to know that Lanaken is situated in the Belgian province with the highest COVID-19 prevalence, and at less than 40 kms from Gangelt, the place of one of the first major outbreaks of COVID-19 in Germany in February 2020.[5] The first patient in the WZC was diagnosed with COVID-19 on February 24^th^and the outbreak was traced to a carnival activity on February 15^th^. The first staff member of wzc Bessemerberg, a nurse, tested positive on PCR on March 20^th^ 2020 and was immediately quarantined at home. The first resident tested positive on polymerase chain reaction (PCR) on 30.3.2020, and most likely was infected much earlier by family. Between March 30^th^ and April 16^th^ 15 (12%) residents died, of which 14 tested positive with PCR, and one tested negative. To manage the outbreak, at April 20^th^ we opened a separate COVID ward within our institution, with a capacity of 22 beds.

As the evolution of the epidemiological picture began to take shape and test results became available, we tried to answer the following questions:

1. What is the prevalence of COVID-19, based on PCR test results in residents and nursing staff?
2. What is the prevalence of IgM and IgG antibodies in residents and nursing staff two weeks after the first resident was diagnosed with COVID-19?
3. What is the influence of age, sex and ward allocation on antibody and PCR tests?
4. What is the relation between PCR and antibody test results, both cross-sectionally and longitudinally?

## Methods

### Testing & outcomes

Before 16.4.2020 both residents and staff members were systematically tested by PCR in case of signs and symptoms suggestive for COVID-19 at the initiative of the general practitioner (GP) or the local coordinating GP of the institution (CRA). The first PCR tests were performed by local laboratories in accordance with the WHO guidelines on nasopharyngeal swabs sampled by the GP or the CRA. On April 16^th^ 2020, all residents and staff members not already tested before were sampled bilaterally in the back of the throat and to the fore of the nose and tested with PCR within the framework of a national governmental survey of nursing homes. A participant was considered PCR+ if he or she tested positive between March 20^th^ and April 16^th^.

Antibody testing was performed by the CRAs, aided by volunteers within the framework of a survey performed to support clinical work between 14 and 16.4.2020. Testing was performed using a point of care lateral flow assay with capillary whole blood from a fingerprick according to the manufacturer’s instructions. We used the Sure Screen Diagnostics Covid-19 IgG/IgM Rapid test casette (Sure Screen Diagnostics, Derby, UK) (Figure 1). Validations by the manufacturer and at the University Hospitals Leuven found a specificity of 95-99%, and a sensitivity of 85-100% for IgG 14-25 days after onset of symptoms. For the IgM, sensitivity was 92% and specificity 99% (instructions of the producer, Sure Screen Diagnostics, Derby, UK).

**Figure 1:**
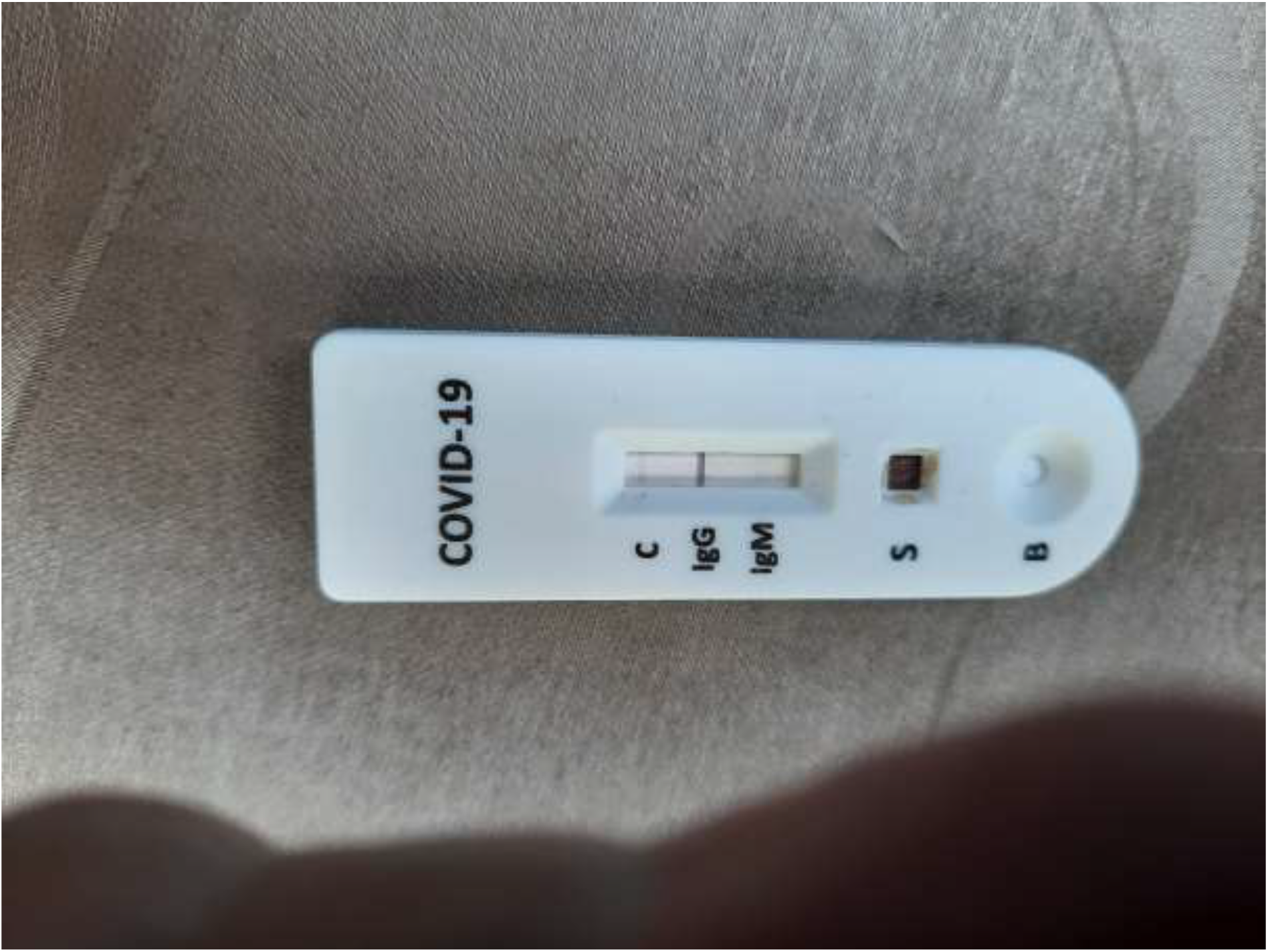
Sure Screen Diagnostics Covid-19 IgG/IgM Rapid test casette

A test was considered positive for IgM/IgG (IgM/IgG +) if at least IgM or IgG was positive. There was only one device failure (the control line was not visible).

Within wzc Bessemerberg, there are under normal circumstances three wards wit 50, 40 and 40 beds respectively. Nursing staff tends to be affiliated to one of these wards. When analysing the association with wards (residents and staff) and working situation (staff), we combined as ‘paramedical’, those staff members who have direct contact with resident, but are not directly working at one specific ward (e.g., physiotherapists, nurses working night shifts, etc.). As ‘nonmedical’ we categorised staff members without regular direct patient contacts (e.g., administration or maintenance).

As soon as the COVID-19 ward was initiated, all PCR positive residents were brought to this ward, unless they were already considered preterminal. A resident was returned to his or her normal room two weeks after the initial positive PCR test, provided that the resident was clinically stable and asymptomatic for at least three days. [6]

All participant-related information was coded and treated according to the Belgian Privacy law and the GDPR. The chairman of the ethical review board of the ZOL hospital in Genk confirmed that a formal ethical review was not required.

### Patient and public involvement

Patients and the public were not directly involved in the design or the recruitment of the study.

### Statistics

All questions were answered in a descriptive way, using bivariate statistical tests (chi^2^ test or oneway ANOVA) or Mantel-Haenszel stratified analysis if appropriate.

To answer the longitudinal questions, we also performed multiple logistic regression, testing the influence of PCR results, participant type (resident or staff), ward, and gender on the presence of sars-CoV-2 antibodies (IgM, IgG, either one of them). Age was not used in the model, because of the overlap between age and role (resident or staff).

## Results

One hundred-thirty residents and 112 staff members were considered for PCR and antibody testing. Some residents were not tested because at the time of the sampling they were either in hospital or deceased. Some staff members decided not to participate or were absent at the time of the sampling. In total, N=119 residents and N=93 staff members were tested. The number of residents and staff members who were tested (denominators) with PCR and with the antibody test are not identical as both tests were not performed in parallel on the same day and not everyone was tested with both tests.

In total, 200 people had a PCR test and 51 of the participants (26%) had a positive test. Of the 118 residents who had a PCR test, 40 (34%) had a positive result. Of the 82 staff members, 11 (13%) had a positive result. Of 188 people tested for antibodies (residents and staff combined), 13% was positive on IgM, 15% on IgG and 19% on at least one of both. Of the residents, 17% tested positive for IgM/IgG (13% for IgM and 15% for IgG). Of the staff members, 20% tested positive for IgM/IgG, 13% for IgM and 16% for IgG (table 1).

**Table 1:** COVID-antibody and PCR test results in residents and staff: numbers (percentages)

|  | IgM + | IgG + | IgM/IgG + | PCR + |
| --- | --- | --- | --- | --- |
| Residents | 13/100 (13%) | 15/100 (15%) | 17/100 (17%) | 40/118 (34%) |
| Staff | 11/88 (13%) | 14/88 (16%) | 18/88 (20%) | 11/82 (13%) |
| Total | 24/188 (13%) | 29/188 (15%) | 35/188 (19%) | 51/200 (26%) |

\* One resident didn't do the PCR test; 19 residents didn't do the IgM/IgG test. 11 members of the staff didn't do the PCR test and 5 staff members didn't do the IgM/IgG test.

### Specification according to the wards, gender, age, and symptoms

Prevalence of antibodies was strongly related to the allocated ward where a resident lives. It was very low on ward 1, and much higher on ward 2 and 3 (table 2).

**Table 2:** COVID-antibody test results according to the wards (residents).

|  | IgM +<br>N (%) | IgG +<br>N= (%) | IgM/IgG + +<br>N (%) | PCR +<br>N (%) |
| --- | --- | --- | --- | --- |
| Ward 1 | 1/40 (3%) | 2/40 (5%) | 2/40 (5%) | 11/51 (22%) |
| Ward 2 | 7/28 (25%) | 7 /28(25%) | 8/28 (29%) | 19/34 (56%) |
| Ward 3 | 5/32 (16%) | 6 /32(19%) | 7/32 (22%) | 10/33 (30%) |
| Total | 13/100 (13%) | 15/100 (15%) | 17/100 (17%) | 40/118 (34%) |

**Table 3:** COVID-antibody test results according to the wards / working place (staff).

|  | IgM +<br>N (%) | IgG +<br>N (%) | IgM or IgG +<br>N (%) | PCR +<br>N (%) |
| --- | --- | --- | --- | --- |
| Ward 1 | 4/22 (18%) | 5/22 (23%) | 7/22 (32%) | 3/20 (15%) |
| Ward 2 | 3/19 (16%) | 3 /19(16%) | 4/19 (21%) | 2/16 (13%) |
| Ward 3 | 4/28 (14%) | 4/28 (14%) | 5/28 (18%) | 3/28 (11%) |
| Paramedicals | 0/10 (0%) | 1/10(10%) | 1/10 (10%) | 1/11 (9%) |
| Other | 0/9 (0%) | 1/9 (11%) | 1/9 (11%) | 2/7 (29%) |
| Total | 11/88 (13%) | 14/88 (16%) | 18/88 (20%) | 11/82 (13%) |

The association between ward or working place and antibody prevalence in staff members is weaker compared to residents (table 3).

No association was found between either IgM or IgG or both and gender, neither in residents nor staff: adjusted odds ratio according to Mantel-Haenszel = 0.59 (0.19-1.83); p = 0.36.

No association was found between either IgM or IgG or both and age, neither in residents nor staff: p = between 0.34 – 0.91 (ANOVA)

There was no significant association between a positive IgM, IgG of one of these and presence of serious symptoms at the time of sampling (OR = 1.17, 1.75,1.4; p = between 0.11 – 0.45).

For all antibodies, a multiple logistic regression analysis resulted in a model with only a positive PCR test as a significant predictor: coefficient = between 1.08 – 2.14; p = between < 0.001 – 0.09).

Gender, ward, and role remained insignificant

### PCR result and the presence of antibodies

We had both PCR and antibody results of 176 participants (99 residents and 77 staff). Of the 42 participants who were positive with PCR, 19 (45%) also had antibodies (IgM/IgG +). Of 134 PCR participants who were negative with PCR, 15 (11%) had antibodies (IgM/IgG +).

In PCR positive residents, the percentage of IgM positive, IgG positive, or at least one of both was 28%, 34%, and 41%. In PCR positive staff, we found 30%, 60%, and 60% (table 4).

**Table 4:**
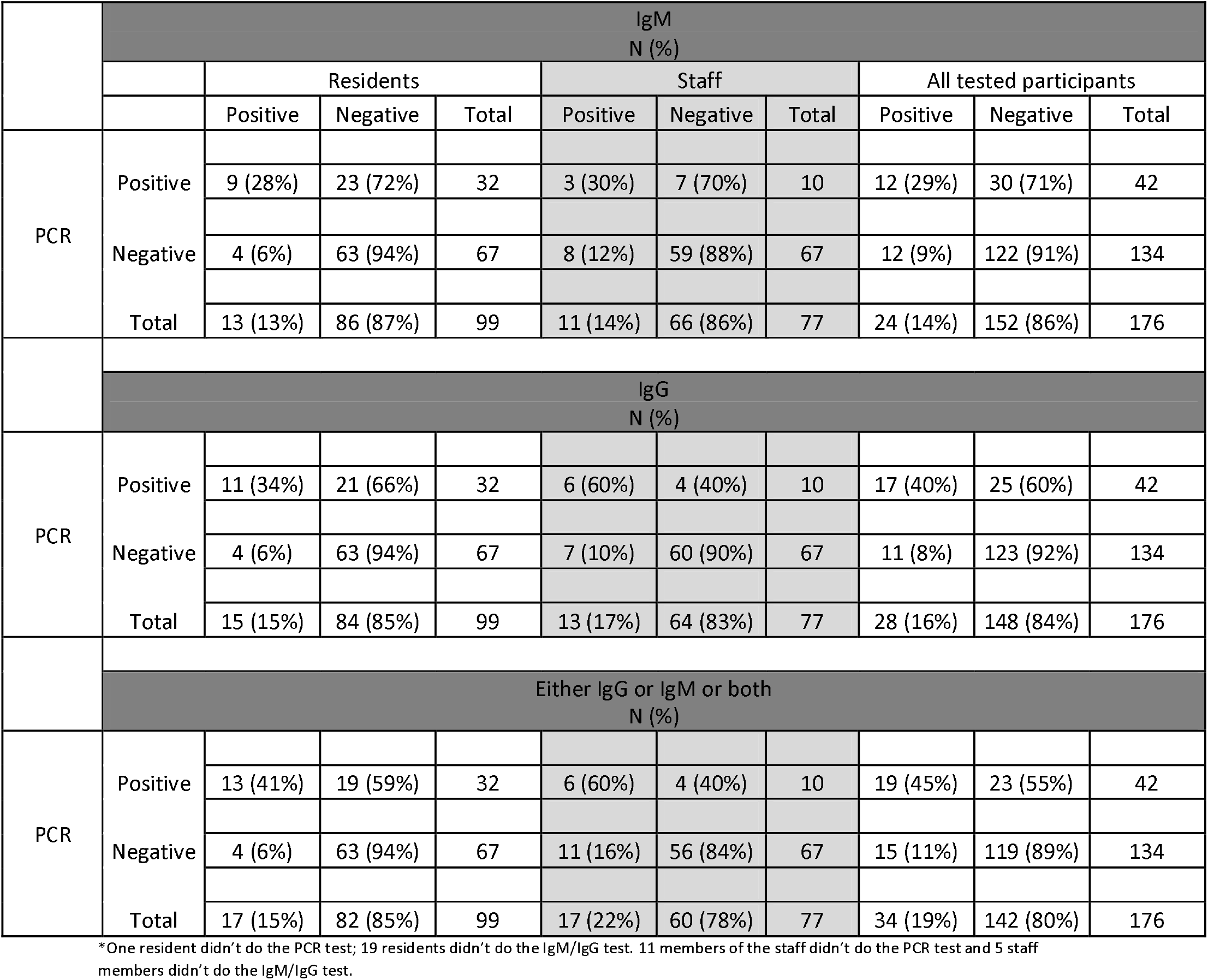
Presence of antibodies and PCR results at the same moment (cross-sectional analysis)

In 16 residents, a positive PCR test was detected before 3.4.2020. Of those, seven were in hospital or deceased on 16.4.2020. Therefore, nine residents of this group were available for antibody testing. They were tested for antibodies between day 11 and 14 after the positive PCR test.

Of those, 4 (44%), 6 (67%) and 7 (78%) tested positive on IgM, IgG, or at least one of them. The positive IgM results were detected on day 12 and 13 after the positive PCR test, the positive IgG results on day 11, 12, 13 and 14. The two IgM/IgG negative tests were performed 14 and 22 days after the positive PCR. When retesting antibodies three weeks later (12.5.2020), both remaining residents also tested positive on IgG.

## Discussion

End of April 2020 the prevalence of IgG antibodies in the general population in Belgium was 3-4% (measured using the Eurimmun IgG Elisa, based on rest-samples of routine blood testing in ambulatory medicine.[7] Recently, the local hospital surveyed its staff and reported presence of COVID antibodies in 6%. Our study reports a presence of IgM and/or IgG in 17% of residents and 20% of nursing staff in a nursing home. The large difference between hospital staff (6%) and nursing home staff (21%) probably relates to the complete absence of good quality protection for nursing home personnel in the first weeks of the outbreak. The prevalence in our participants is much higher compared to the general population, although even this is largely insufficient for herd immunity, now or in the near future.

The current gold standard for the diagnosis of COVID-19 is the detection of viral RNA in respiratory tract samples. Its sensitivity is, however, only around 54%-74% for nasopharyngeal swabs. False-negative results can occur, due to sampling error and in patients with low viral loads, especially in patients who present at day 8 after infection or later, and mild cases.[8] In our group of PCR negative participants this shows as 11% (15/134) having a positive IgG or IgM or both.

Antibodies can be detected within two weeks after detection of a positive PCR. In our group, it was 44% for IgM and 78% for either IgG or IgM). As also found elsewhere [9], the moment at which antibodies can be detected with a high sensitivity is therefore insufficient for their use as an early diagnostic test. Out of a group of nine residents, who were tested some two weeks after a positive PCR test, two were IgM and IgG negative. When retesting three weeks later, however, both showed positive IgGs, suggesting that at least in this small group all PCR positive residents formed IgG antibodies, if given a sufficient amount of time.

In PCR positive participants, presence of either IgG or IgM was found in 41% of residents and 60 % of nursing staff. This difference suggests that antibody production could be lower or slower in residents of nursing homes, although we cannot rule out that PCR-positive staff members were on average infected earlier than residents

Presence of antibodies in residents differed largely according to the wards where they are living. It is minimal at the first ward and far more frequent at ward two and three (5%, 29% and 22% respectively of the residents have IgM or IgG antibodies). These are also the two wards where outbreak effects became first clear, and which showed the largest numbers of illnesses and deaths. These differences were far less in staff members. The large differences between wards in the number of residents support the emerging plans to reorganise homes for the elderly in smaller living units, i.e. with ten rooms instead of 30 or 40 as is currently the standard in Belgium.

### Strengths and limitations

Testing and reading the results was easy to perform. It was also reassuring that – with one exception – the internal procedural control was positive in all tests, which suggests correct sampling and testing. The one exception was one of the first tests performed, probably due to an insufficient amount of blood. Other validation studies resulted in sensitivities after 14-17 days between 85-100 % (instructions of the producer, Sure Screen Diagnostics, Derby, UK). This is less than perfect, but similar to common Elisa tests. Specificity, which in this situation is the most important, was between 96-99%; which is excellent.

Some eligible people were not tested. For staff, this mostly resulted from absence of work without any direct association with COVID-related disease. We therefore do not expect any selection bias resulting from this incompleteness of data. For residents, however, absence resulted mostly from hospitalisation or death. Of course, this can be related to presence of COVID-related disease and some degree of selection bias should be assumed. These numbers are low (mostly less than ten), however, and will not largely influence our results or conclusions.

### Consequences of our study for clinical work and for future research

Information on antibody status can be useful to support decisions in the further course after a COVID infection. For residents, presence of IgG in association with absence of signs and symptoms and a period of at least two weeks after the first positive PCR result, could be considered a criterion for discharge from the COVID ward to the normal rooms. A negative PCR result cannot be considered the ultimate criterium for discharge since PCR can be both false-negative later during the course of the infection and false-positive due to detection of residual RNA after viral clearance. For staff members, the presence of IgG means they are unlikely to get reinfected in the next months and spread the virus. Staffing our nursing homes with staff who tested positive for IgG could help bring down the high case-fatality rates. [10]. It is not sure, however, how long immunity, if present, will last.

Antibody testing can also be used for epidemiological purposes. Follow-up of a population with antibody testing describes the gravity and evolution of the epidemic. [6, 11, 12] It can also show how far away we still are from herd immunity. Finally, it can help to identify convalescent cases or people with milder disease who might have been missed by a PCR survey when performing network analysis [13].

It looks appropriate, to follow the presence of antibodies over time in (a subgroup of) our antibody-positive participants. After 6, 12, 24, and 60 months we can then study if IgG antibodies are still present.

## Data Availability

dates are available on reasonable request

## Acknowledgements

We thank all participant residents and staff of wzc Bessemerberg, as well as all volunteers who helped with organising and sampling.

## Authors Contribution

All authors attest they meet the ICMJE criteria for authorship. All authors have contributed to, seen and approved the final version of the manuscript.

BF, VP, VRM designed the research. BF, CP, GM conducted the research. BF, CP, GM, JV, JDL, MVDE, VP, VRM analysed the data. BF, CP, GM, JV, JDL, MVDE, VP, VRM prepared and discussed the manuscript.

## A patient consent form

not required as confirmed by the chairman if the ethical review board of the ZOL hospital in Genk (letter dd 11.5.2020)

## Data availability statement

No additional data are available.

## References

1. Haveri, A., Smura, T., Kuivanen, S. et al., Serological and molecular findings during SARS-CoV-2 infection: the first case study in Finland, January to February 2020. Euro Surveill, 2020. 25(11).

2. WHO, WHO Director-General’s opening remarks at the media briefing on COVID-19 -11 March 2020. 2020.

3. Spiteri, G., Fielding, J., Diercke, M. et al., First cases of coronavirus disease 2019 (COVID-19) in the WHO European Region, 24 January to 21 February 2020. Euro surveillance: bulletin Europeen sur les maladies transmissibles = European communicable disease bulletin, 2020. 25(9): p. 2000178.

4. Sciensano, COVID-19—EPIDEMIOLOGISCH BULLETIN VAN 3 MEI2020. https://covid-19.sciensano.be/sites/default/files/Covidl9/covid-19_daily_report_20200503_-_nl.pdf/, 2020. (last consulted May 5th)

5. Streeck, H., Exner, M., Vorläufiges Ergebnis und Schlussfolgerungen der COVID-19 Case-Cluster-Study (Gemeinde Gangelt). Preliminary Report of the Covid-19 Case-Cluster-Study to the Government of North Rhine-Westphalia(09.04. 2020), accessed on, 2020. 9: p. 2020.

6. Inglesby, T.V., Public Health Measures and the Reproduction Number of SARS-CoV-2. JAMA, 2020.

7. Finoulst, M., Kunnen we groepsimmuniteit tegen covid-19 alleen met een vaccin sneller bereiken? https://www.gezondheidenwetenschap.be/. 2020. (last consulted May 5th)

8. Yang, Y., Yang, M., Shen, C. et al., Laboratory diagnosis and monitoring the viral shedding of 2019-nCoV infections. MedRxiv, 2020.

9. Abbasi, J., The Promise and Peril of Antibody Testing for COVID-19. JAMA, 2020.

10. Del Rio, C. and P.N. Malani, COVID-19—new insights on a rapidly changing epidemic. Jama, 2020.

11. Gandhi, M., D.S. Yokoe, Havlir, D.V., Asymptomatic Transmission, the Achilles’ Heel of Current Strategies to Control Covid-19. 2020, Mass Medical Soc.

12. Kissler, S.M., Tedijanto, C., Goldstein, E., et al., Projecting the transmission dynamics of SARS-CoV-2 through the postpandemic period. Science, 2020.

13. Yong, S.E.F., Anderson, D.E., Wei, W., et al., Connecting clusters of COVID-19: an epidemiological and serological investigation. The Lancet Infectious Diseases, 2020.

